# The COVID-19 pandemic shifted the Veterans Affairs System toward being a payer and virtual care provider: is it sustainable?

**DOI:** 10.1101/2021.05.31.21258031

**Authors:** Liam Rose, Linda Diem Tran, Steven M. Asch, Anita Vashi

**Affiliations:** Health Economics Resource Center, VA Palo Alto Health Care System; Stanford Surgery Policy Improvement and Education Center, Stanford Medicine; Center for Innovation to Implementation, VA Palo Alto Health Care System; Division of Primary Care and Population Health, Stanford Medicine; Center for Innovation to Implementation, VA Palo Alto Health Care System; Department of Emergency Medicine, University of California, San Francisco

**Keywords:** Veterans Health, United States Department of Veterans Affairs, Telemedicine, Access to Health Care, Health Care Economics and Organizations

## Abstract

**Objective:** To examine how VA shifted care delivery methods one year into the pandemic.

**Study Setting:** All encounters paid or provided by VA between January 1, 2019 and February 27, 2021.

**Study Design:** We aggregated all VA paid or provided encounters and classified them into community (non-VA) acute and non-acute visits, VA acute and non-acute visits, and VA virtual visits. We then compared the number of encounters by week over time to pre-pandemic levels.

**Data Extraction Methods:** Aggregation of administrative VA claims and health records.

**Principal Findings:** VA has experienced a dramatic and persistent shift to providing virtual care and purchasing care from non-VA providers. Before the pandemic, a majority (63%) of VA care was provided in-person at a VA facility. One year into the pandemic, in-person care at VA’s constituted just 33% of all visits. Most of the difference made up by large expansions of virtual care; total VA provided visits (in person and virtual) declined (4.9 million to 4.2 million) while total visits of all types declined only 3.5%. Community provided visits exceeded prepandemic levels (2.3 million to 2.9 million, +26%).

**Conclusion:** Unlike private health care, VA has resumed in-person care slowly at its own facilities, and more rapidly in purchased care with different financial incentives a likely driver. The very large expansion of virtual care nearly made up the difference. With a widespread physical presence across the U.S., this has important implications for access to care and future allocation of medical personnel, facilities, and resources.

## Introduction

The Veterans Health Administration (VA) manages a nearly $100 billion per year integrated health care system for over 9 million enrollees.^1^ This makes it the third largest federal outlay on healthcare behind Medicare and Medicaid. VA facilities span every state (as well as Puerto Rico, the Virgin Islands, Guam, American Samoa, and the Philippines). In addition, in the wake of concerns about access, the VA has increased the amount of care it purchases from community providers.

A multitude of factors make it difficult to predict veterans’ health care needs and ensure access meets demand. First, veteran demographics are changing. Despite a three-decade long decline in the size of the veteran population, the number of VA patients has been increasing.^1^ Policy and eligibility changes also impact the number veterans eligible for VA health care benefits. Moreover, many veterans who are eligible for VA health care have other insurance coverage and may only rely on VA to meet some of their health care needs. This reliance on VA can fluctuate based on economic conditions, U.S. health care policy changes, and location. To provide Veterans with more flexibility, the VA has increased spending on community care. From 2014 to 2018, the community care budget increased 82% and comprised nearly 20% of VA’s health care budget.^2^

Like other systems, VA has faced unprecedented challenges responding to the COVID-19 pandemic. While VA’s large size, diverse operating environments, and geographically dispersed patient population make it difficult to pivot nimbly, VA was able to leverage its existing infrastructure and prior planning to rapidly scale existing virtual care services - a key component of VA’s response to the pandemic. In this study, we examine how VA care patterns shifted in response to the COVID-19 pandemic. Our analysis aims to show how care volumes and delivery methods evolved over the subsequent year to better understand how the organization and delivery of VA health care can be tailored to meet the needs of Veterans in the short-term without becoming inefficient in the long run.

## Methods

To capture all health care encounters in VA and those paid for by VA in the community, distinct VA and Community Care encounters from January 2019 to February 2021 were extracted from national VA administrative databases housed in the VA Corporate Data Warehouse.

Encounters were then classified into mutually exclusive categories by the type and location of care delivered. VA acute encounters included VA emergency department (ED) and urgent care (UC) visits and acute inpatient hospital days. For inpatient stays lasting multiple days, a visit was counted for each day of the stay. Multiple ED visits on the same day were only counted once. Community acute care visits included ED and acute inpatient hospital days provided in community settings. Remaining VA encounters (outpatient care, rehabilitation care, ancillary and diagnostic encounters) were further categorized as in-person care or virtual care (telephone or video-based, not including secure messaging). Similarly, remaining Community Care visits included all non-acute encounters. However, Community Care visits were not further categorized as in-patient or virtual but both types were included in totals, as virtual community encounters were less than 0.1% of all non-acute community encounters.

Encounter data was aggregated by epidemiological weeks that start on the first Sunday of each calendar year. To estimate total volume of missing visits in 2020, we performed kernel-weighted local polynomial regression of total weekly visits on the numeric week of the year in 2019 and applied the smoothed values -- expected total visits per week had 2019 levels persisted -- to the numeric weeks in 2020. We subtracted the smoothed values or totals from actual 2020 totals and summed the differences across all 52 weeks.

Limitations of this study include that there is likely a lag in processing, reporting, and adjudication of more recent community care claims. However, this means that current estimates of Community Care encounters are an underestimate. Also, due to a limitation in available claims data, we were unable to reliably categorize Community Care as in-person or virtual.

## Results

VA provided or paid for 188.3 million encounters between January 2019 and February 2021. **Figure 1** provides the volume of encounters over time by type and location. As expected, overall utilization dropped precipitously in March and April of 2020, coinciding with the start of the COVID-19 pandemic. However, virtual care in VA expanded swiftly. Strikingly, total encounters have yet to recover to either 2019 or early 2020 pre-pandemic levels, with 2019 levels shown with a dashed line. The estimated total volume of missing encounters relative to 2019 is 13.2 million. Note that this is a conservative estimate given strong yearly growth in encounter volume prior to the pandemic. **Figure 2** shows these same data in proportions to better illustrate the change in the distribution of care categories over time.

**Figure 1:**
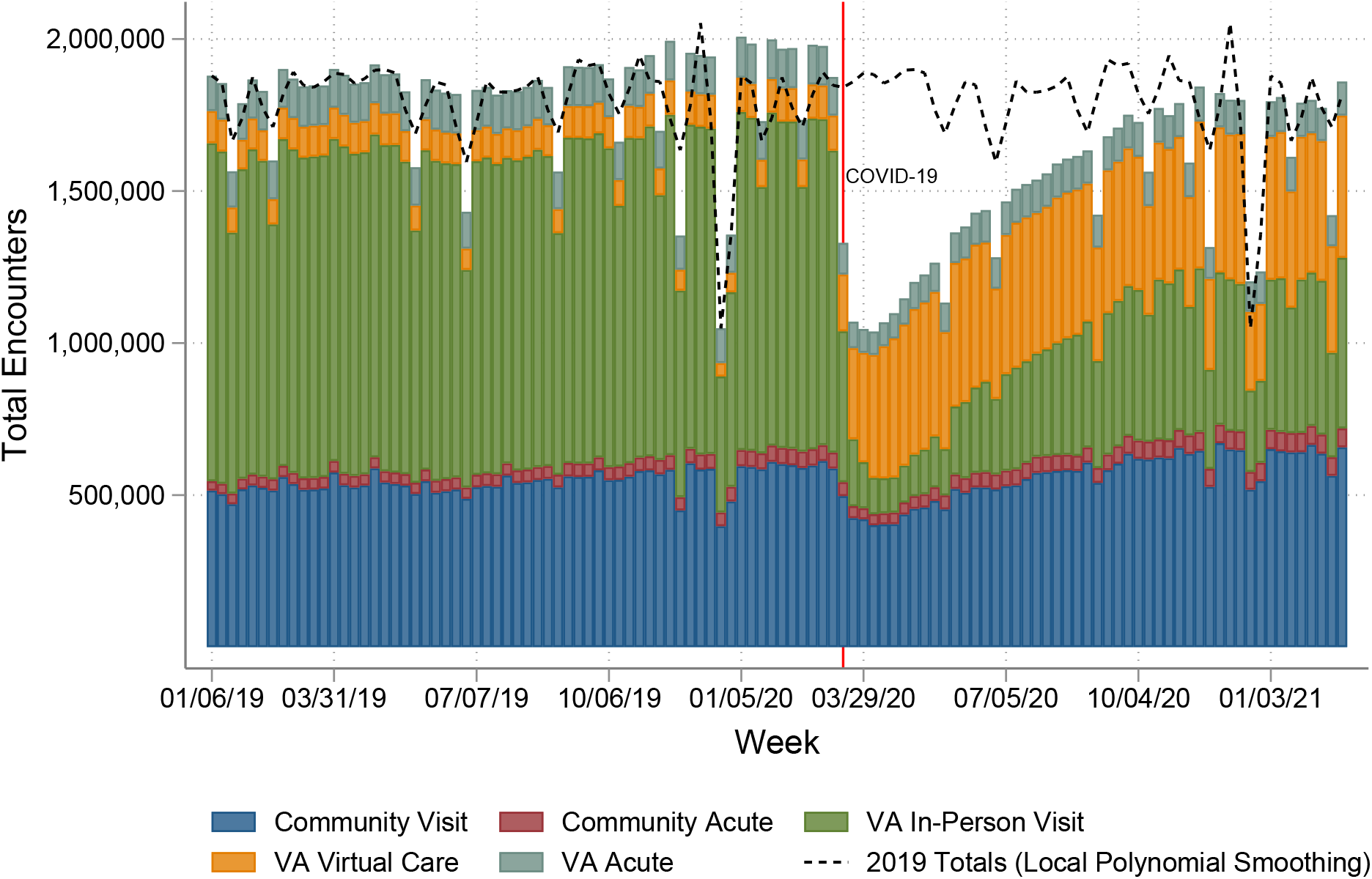
All VA and Community Care encounters, January 2019-Febuary 2021. Notes: All VA provided visits (VA In-Person Visit, VA Virtual Care Visit, and VA Acute) along with all VA paid visits (Community Visit and Community Acute) from January 2019 to February 27, 2021. Acute refers to inpatient, emergency department, and urgent care visits. VA virtual care refers to both telephone and video visits. The dashed lined represents a smoothed local polynomial over 2019 data.

**Figure 2:**
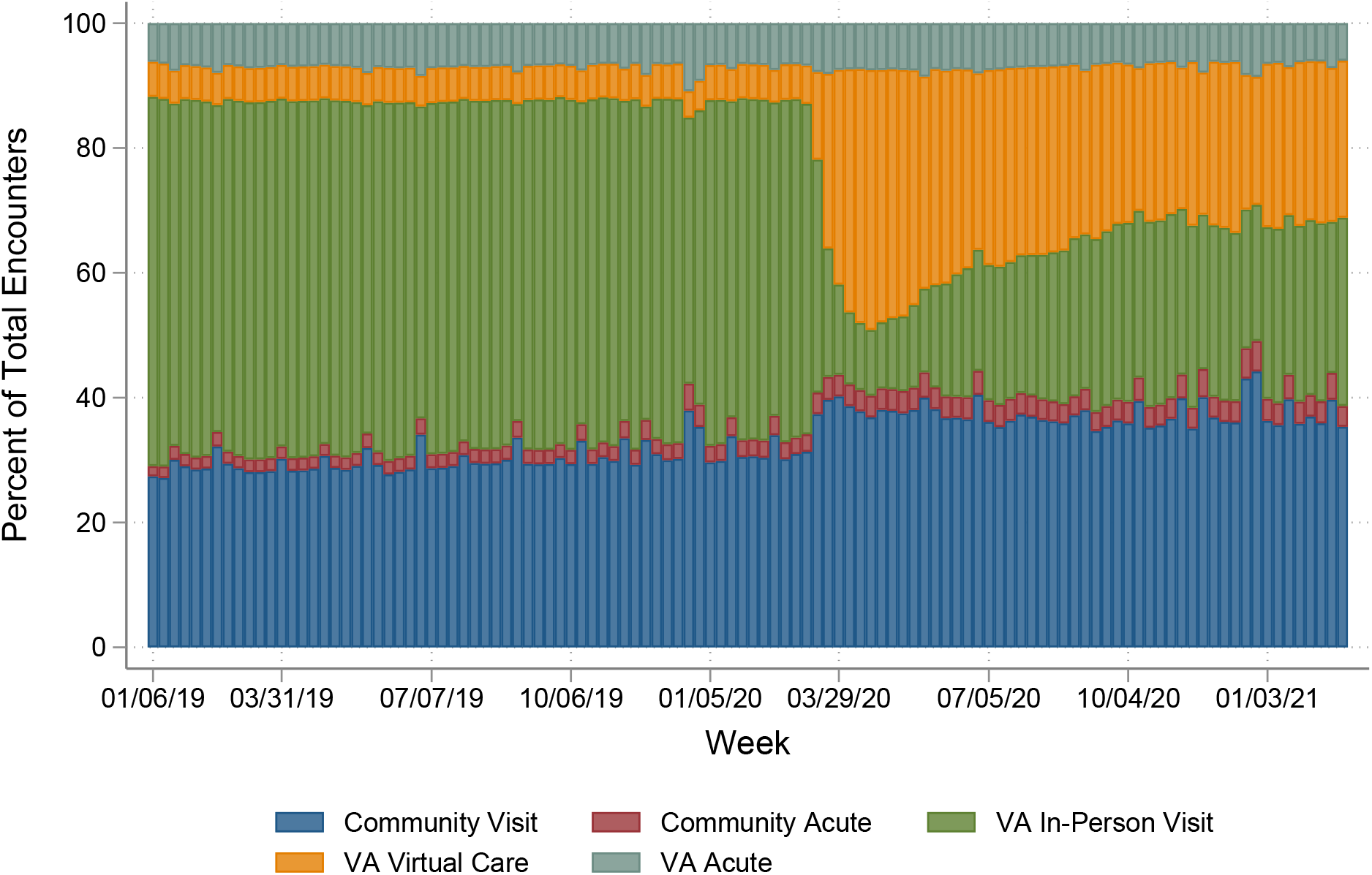
Distribution of encounters by visit location and type. Notes: Breakdown of all VA provided and paid visits from January 2019 to February 27, 2021. Acute refers to inpatient, emergency department, and urgent care visits. VA virtual care refers to both telephone and video visits.

**Table 1** describes the share of each category of care in April 2019, April 2020, December 2019, and December 2020 along with the percent change. Virtual care in VA increased from 6% (n=454,152) in April 2019 to 40% (n=1,894,878) in April 2020 before falling to 26% (n=1,863,417) in December 2020. Non-acute Community Care increased from 28% in April 2019 to 41% in April 2020 but held nearly steady through the end of the year (n=2,374,314 in April 2019 to n=2,712,899 in December 2020). As of December 2020, VA in-person care constituted just 33% of VA paid or provided care.

**Table 1:** Change in VA and Community Care encounters during COVID-19 and relative to 2019.

|  | April 2019 |  | April 2020 |  | % Change | December 2019 |  | December 2020 |  | % Change |
| --- | --- | --- | --- | --- | --- | --- | --- | --- | --- | --- |
|  | Total | % | Total | % |  | Total | % | Total | % |  |
| **Total Encounters** | 8248675 |  | 4722492 |  | -42.7% | 7497813 |  | 7335080 |  | -2.2% |
| **Community Visits** | 2375460 | 28.8 | 1792779 | 37.9 | -24.5% | 2376873 | 31.7 | 2798822 | 37.5 | 17.8% |
| **Community Acute** | 161139 | 1.9 | 159600 | 3.3 | -1.0% | 212199 | 2.8 | 269752 | 3.5 | 27.1% |
| **VA In-person** | 4703443 | 57.0 | 531676 | 11.3 | -88.7% | 3957218 | 52.7 | 1916660 | 26.5 | -51.6% |
| **VA Acute** | 554491 | 6.7 | 343572 | 7.3 | -38.0% | 549380 | 7.3 | 487080 | 6.7 | -11.3% |
| **VA Virtual Care** | 454142 | 5.5 | 1894865 | 40.2 | 317.2% | 402143 | 5.3 | 1862766 | 25.8 | 363.2% |

*Source: Author analysis of VA data. Notes: Proportion in April 2019 gives the proportion of all VA paid and provided visits in that category in April 2019, and Proportion in April 2020 and Proportion in December 2020 gives the same from April 2020 and December 2020 respectively. Percent Change April 2019 – April 2020 and Percent Change December 2019 – December 2020 give the percent change in that visit category.*

## Discussion

This analysis demonstrates how utilization patterns changed for VA enrollees during the COVID-19 pandemic. Like other systems, VA experienced large reductions in care early in the pandemic. However, VA was well-positioned to quickly transition to providing large amounts of virtual care.^5^ More surprisingly, however, VA in-person care declined much more than Community Care and has not yet recovered. As of December 2020, VA virtual care and Community Care made up 67% of total VA paid and provided care.

Our results likely reflect several trends. First, our findings indicate that VA has likely adopted a more conservative reopening strategy, compared to community providers who have different financial incentives to resume in-person care. We found that Community Care paid for by VA returned to pre-pandemic levels by September 2020. This is consistent with private health care systems that have reported to have recovered within 5% of pre-pandemic inpatient and outpatient volume by September 2020.^6,7^ It may be that as VA continues to reopen, the share of in-person VA encounters will steadily rebound over a longer period. However, it is possible that in the interim, the shift toward Community Care may be preferred by veterans and thus semi-permanent. Alternatively, if VA and veterans continue to embrace virtual care, in-person VA visits may never fully rebound.

Some of the shift towards Community Care may reflect VA’s longer-term strategy to increase options for veterans and outsource care to the community as formalized by the Maintaining Systems and Strengthening Integrated Outside Networks Act of 2018 (VA MISSION Act) and its predecessor, the Veterans Access, Choice and Accountability Act of 2014 (VA Choice Act). There are inherent tensions in the goals of purchased care, reflecting uncertainty about the extent to which policymakers wish to preserve VA’s primary function as a health care provider or allow VA to provide more care through the community. In either scenario, a substantial reallocation of VA resources and capabilities will be needed to meet the near-term demand for health services among veterans.

What is clear is that the COVID-19 pandemic has resulted in substantial and persistent shifts in care patterns in the VA health care system that warrant continued monitoring as VA resumes normal operations. Telehealth adoption has been widespread, but existing trends pushing VA toward being a mixed payer and provider seem to have accelerated. VA should be prepared to reallocate resources to care for veterans and will need to articulate a clear strategy for how purchased care should be used and how it fits into VA’s broader health care mission.

## Data Availability

Data used in this manuscript contain personal health information and cannot be released.

